# COVID-19 Prevention Facilitators and Barriers among Specific Ethnic Minority Communities in Rural Ohio

**DOI:** 10.1101/2021.10.21.21265302

**Authors:** Paran Pordell, Hammad Ali, Gisela Medina Martinez, Brandi Taylor, Karthik Kondapally, Ellen Salehi, Sietske de Fijter, Nikki Hayes, Spencer Lloyd

## Abstract

**Objective:** To assess knowledge, beliefs, and behaviors concerning COVID-19 among Guatemalan, Marshallese, and Amish populations in rural Ohio; identify individual, interpersonal, community, and structural level challenges within each community; and provide population-specific recommendations to prevent and mitigate further SARS-CoV-2 transmission among these rural communities.

**Methods:** We conducted 30 key informant interviews in four rural counties in Ohio, in May 2020. Three teams of two investigators conducted interviews with local health department staff, community members, meat packing plant management, and community leaders from three communities disproportionately affected by the COVID-19 pandemic [Guatemalan (N=12), Marshallese (N=7), Amish (N=11)]. We used the Social Ecological Model to identify and categorize themes.

**Results:** Emerging and overall themes were identified and defined. Investigators identified COVID-19 knowledge gaps, myths, and misinformation, food insecurity, community cohesion, stigma, community culture and norms, lack of workplace safety policies, and access to testing as key themes to COVID-19 prevention.

**Conclusions:** Understanding specific barriers and identifying facilitators that most effectively provide resources, healthcare services, education, and social support tailored to specific communities would help deter SARS-CoV-2 transmission.

## Introduction

Infection with Severe Acute Respiratory Syndrome Coronavirus 2 (SARS-CoV-2), the virus that causes Coronavirus Disease (COVID-19), is a significant public health threat and causes substantial morbidity and mortality. COVID-19 has had a disproportionate impact on several groups including older age groups, people with chronic diseases, people who lack access to adequate health care, people from some racial and ethnic minority groups, front-line healthcare workers, and people with immunocompromising conditions (1-4). As of March 4, 2021, the United States reported over 28.5 million COVID-19 cases and approximately 517,224 deaths due to the disease(5). During the same timeframe, Ohio reported 833,772 confirmed cases and 16,750 confirmed deaths from COVID-19 (6). Nationwide, outbreaks generally started in large urban cities and shifted to rural areas, creating a multitude of outbreaks that disproportionately impacted several racial and ethnic minority groups and religious communities (7-9).

While investigating rural hot spots in Ohio in May 2020 and reviewing daily case counts, Marshallese and Guatemalan communities were found to be at increased risk for SARS-CoV-2 infection. Some Marshallese and Guatemalan residents lived near and/or worked in meat and poultry processing plants where outbreaks were occurring. Many plants in rural areas across the country employ larger numbers of individuals from racial or ethnic minority groups (10). Working close to other employees on the processing line for the duration of their shift places plant workers at increased risk for SARS-CoV-2 infection (11). During the same time, we also learned of a reported SARS-CoV-2 outbreak in a rural Amish community in a different part of Ohio.

The Ohio Department of Health (ODH) prioritized these COVID-19 hotspots for further investigation due to requests for assistance from local health departments, current COVID-19 case counts, and the potential risk for further SARS-Co-V-2 transmission to other Ohio rural counties outside of these communities. Some researchers define infectious disease hotspots as geographic areas with higher disease burden (incidence or prevalence) or increased disease transmission efficiency (12). To better understand why outbreaks occurred within specific populations in rural Ohio, we assessed knowledge; beliefs; behaviors; and facilitators and barriers to implementing COVID-19 preventive measures within the home, work, and community environments. We focused on three populations identified by the local and state health departments as experiencing outbreaks of SARS-CoV-2 infections: Rural Ohio residents from Guatemala, the Marshall Islands, and an Amish community.

We used the social ecological model (SEM) as our study framework. The social ecological perspective on health promotion is based on the notion that behavior is shaped by certain factors including: 1) individual factors; 2) interpersonal processes and primary groups; 3) community factors; and 4) societal or structural-level factors (13). The SEM (Figure 1) provides an appropriate theoretical framework for this qualitative study because empowered and sustainable change is necessary at each of these levels to increase knowledge about COVID-19, promote prevention efforts, and reduce health disparities among groups disproportionately impacted by the pandemic (14). Our study objectives included: 1) assessing knowledge, beliefs, and behaviors, concerning COVID-19 among members of these three ethnic minority local communities in rural Ohio; 2) identifying individual, interpersonal, community, and structural-level challenges within each local community; and 3) providing specific recommendations to prevent and mitigate further SARS-CoV-2 transmission in these rural communities.

**Figure 1.**
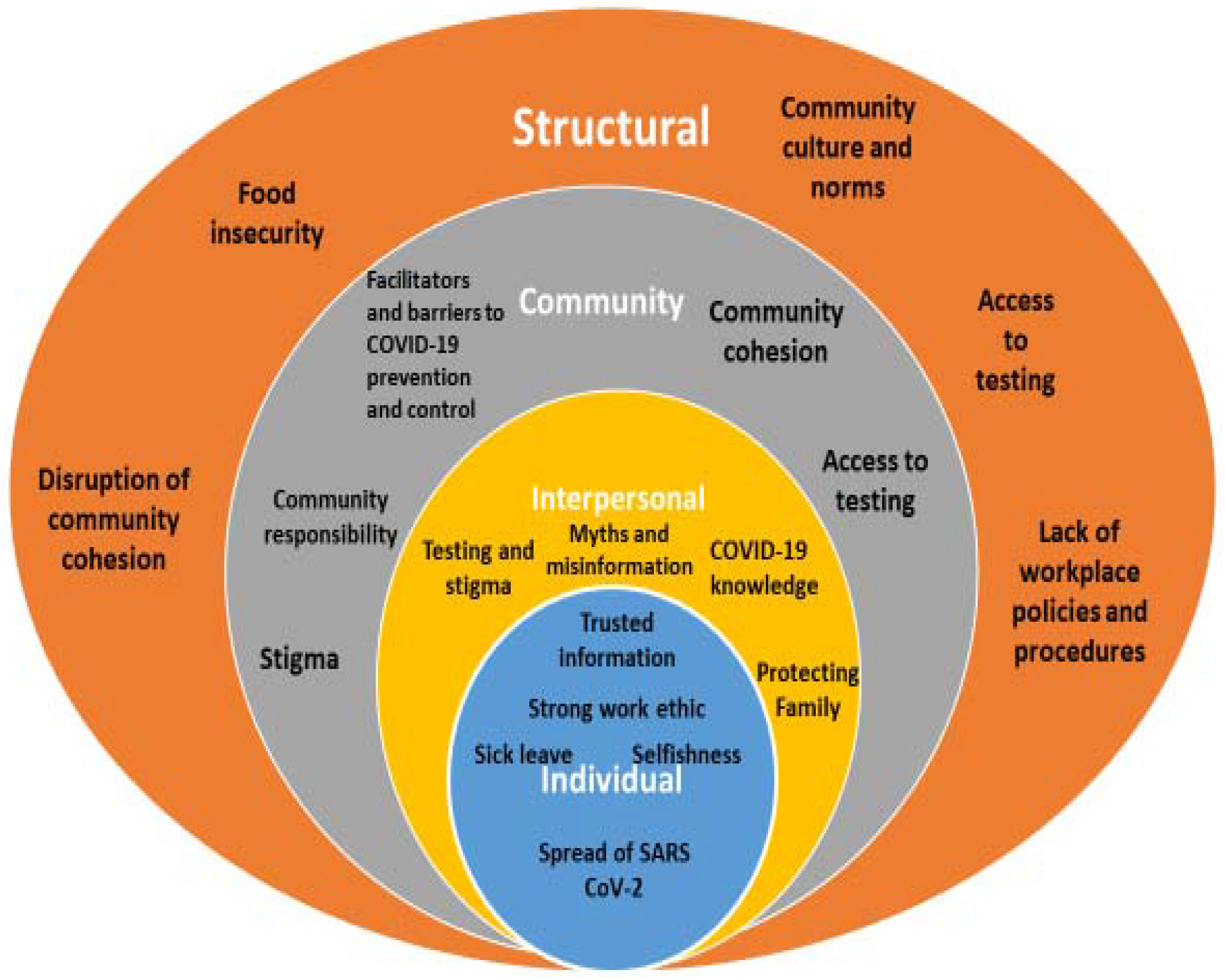
COVID-19 Social Ecological Model and Study Themes^13^

## Materials and methods

The Centers for Disease Control and Prevention (CDC), ODH, several county health departments, and a community-based organization collaborated to conduct thirty key informant interviews (KIIs) with local health department staff, meat processing plant management, and community members and leaders of Marshallese (N=7), Guatemalan (N=12), and Amish (N=11) communities in four rural counties. We used a combination of convenience and snowball sampling (where study participants recruit additional participants from their acquaintances and peers) where the initial interview participants were identified by the ODH and local health departments. We relied on ODH, local health department, community-based organization staff, and community leaders to facilitate community entrance and participation.

We used a semi-structured key informant interview guide to capture community member perspectives and separate, tailored semi-structured guides to conduct community leader and meat processing plant manager interviews. The study team obtained oral informed consent prior to interviews. We pilot-tested the interview guides and revised questions to ensure clarity. Three teams of two investigators (an interviewer and a notetaker) conducted the interviews and both took notes which were transcribed by one investigator from each team. Any discrepancies in the notes were resolved after discussion among team members.

To build trust, respect privacy concerns, and maintain confidentiality among participants, KIIs were not audio-recorded. Interviews were conducted in the languages preferred by participants. Spanish interview responses were transcribed in Spanish and then translated into English by a bilingual study team member. One interview was conducted in English, questions were translated into K’iche’ (Mayan dialect) by an interpreter, and responses were translated and transcribed in English by the interpreter and interviewer, respectively. The study team reflected on their individual biases, backgrounds, and similarities and differences with the study participants before and after interviews and during transcript review as part of the ongoing and fluid research process.

We analyzed the qualitative data manually, using a phenomenological theoretical framework, which focused on understanding the lived experiences of our study populations during the COVID-19 pandemic (15-16). The study team conducted thematic analyses of interview results by ethnic minority population using an iterative process until theme saturation was reached (17-18). Each transcript was reviewed individually by the notetaker and interviewer for that interview, to identify emerging themes, which were categorized in a table that listed emerging themes, overall themes, theme definitions, illustrative quotes to reflect each theme, and relevant summary notes (see Table 1). The study team met to pare down and come to consensus on themes.

**Table 1.**
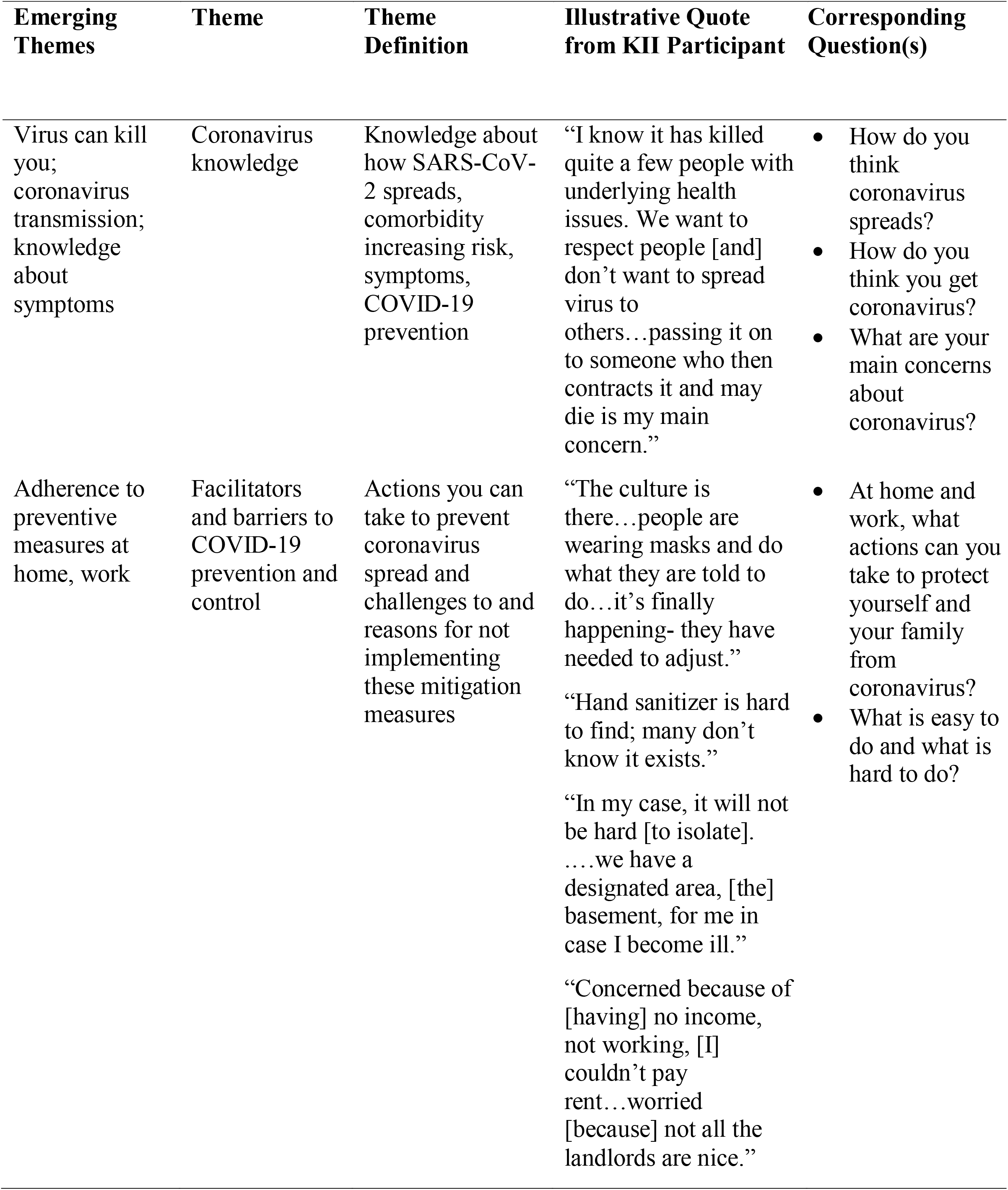
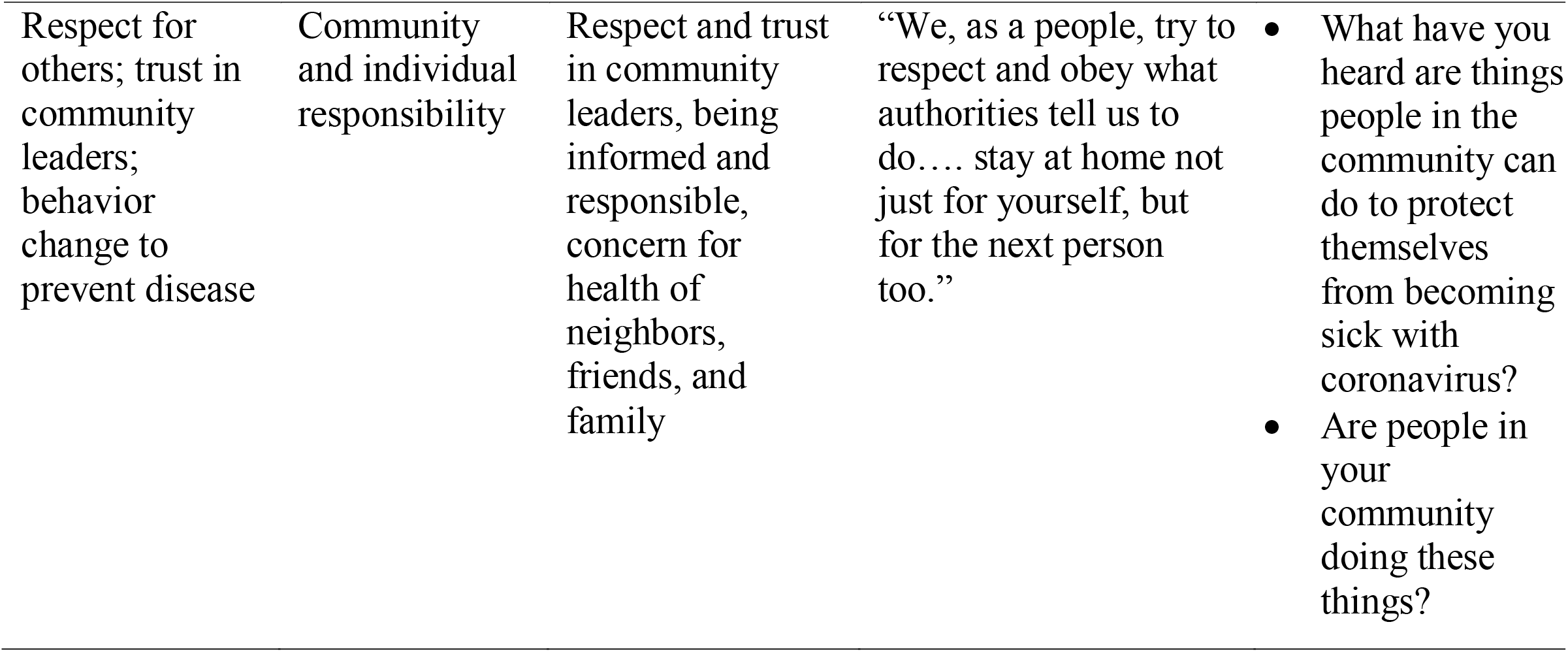
Examples From COVID-19 Study Team Key Informant Interview Themes Table

| **Emerging Themes** | **Theme** | **Theme Definition** | **Illustrative Quote from KII Participant** | **Corresponding Question(s)** |
| --- | --- | --- | --- | --- |
| Virus can kill you; coronavirus transmission; knowledge about symptoms | Coronavirus knowledge | Knowledge about how SARS-CoV-2 spreads, comorbidity increasing risk, symptoms, COVID-19 prevention | “I know it has killed quite a few people with underlying health issues. We want to respect people [and] don’t want to spread virus to others...passing it on to someone who then contracts it and may die is my main concern.” | • How do you think coronavirus spreads?<br>• How do you think you get coronavirus?<br>• What are your main concerns about coronavirus? |
| Adherence to preventive measures at home, work | Facilitators and barriers to COVID-19 prevention and control | Actions you can take to prevent coronavirus spread and challenges to and reasons for not implementing these mitigation measures | “The culture is there...people are wearing masks and do what they are told to do...it’s finally happening- they have needed to adjust.”<br>“Hand sanitizer is hard to find; many don’t know it exists.”<br>“In my case, it will not be hard [to isolate]. ....we have a designated area, [the] basement, for me in case I become ill.”<br>“Concerned because of [having] no income, not working, [I] couldn’t pay rent...worried [because] not all the landlords are nice.” | • At home and work, what actions can you take to protect yourself and your family from coronavirus?<br>• What is easy to do and what is hard to do? |
| Respect for others; trust in community leaders; behavior change to prevent disease | Community and individual responsibility | Respect and trust in community leaders, being informed and responsible, concern for health of neighbors, friends, and family | “We, as a people, try to respect and obey what authorities tell us to do.... stay at home not just for yourself, but for the next person too.” | • What have you heard are things people in the community can do to protect themselves from becoming sick with coronavirus?<br>• Are people in your community doing these things? |

Emerging themes were collapsed into overall themes and a definition for each theme was created. All transcribed notes, data analyses, and study files were stored on password-protected computers, and only the study team had access to the data. The study was reviewed by the CDC ethics committee and approved as non-research. It was conducted consistent with applicable federal law and CDC policy.^i^

## Results

During our in-depth interviews with community members and leaders we uncovered themes corresponding to and impacting all SEM levels (Individual, Interpersonal, Community, and Structural) (Figure 1). Some themes matched to multiple SEM categories and although one objective of the study was to identify and define themes by ethnic minority population, we also identified common themes shared by some or all three populations (see Table 2). For example, access to testing for all three communities was identified as a challenge at community and structural levels and community cohesion was identified as a strength or concern at interpersonal and community levels within Marshallese and Amish communities because these themes impacted participant experiences at multiple SEM levels.

**Table 2.**
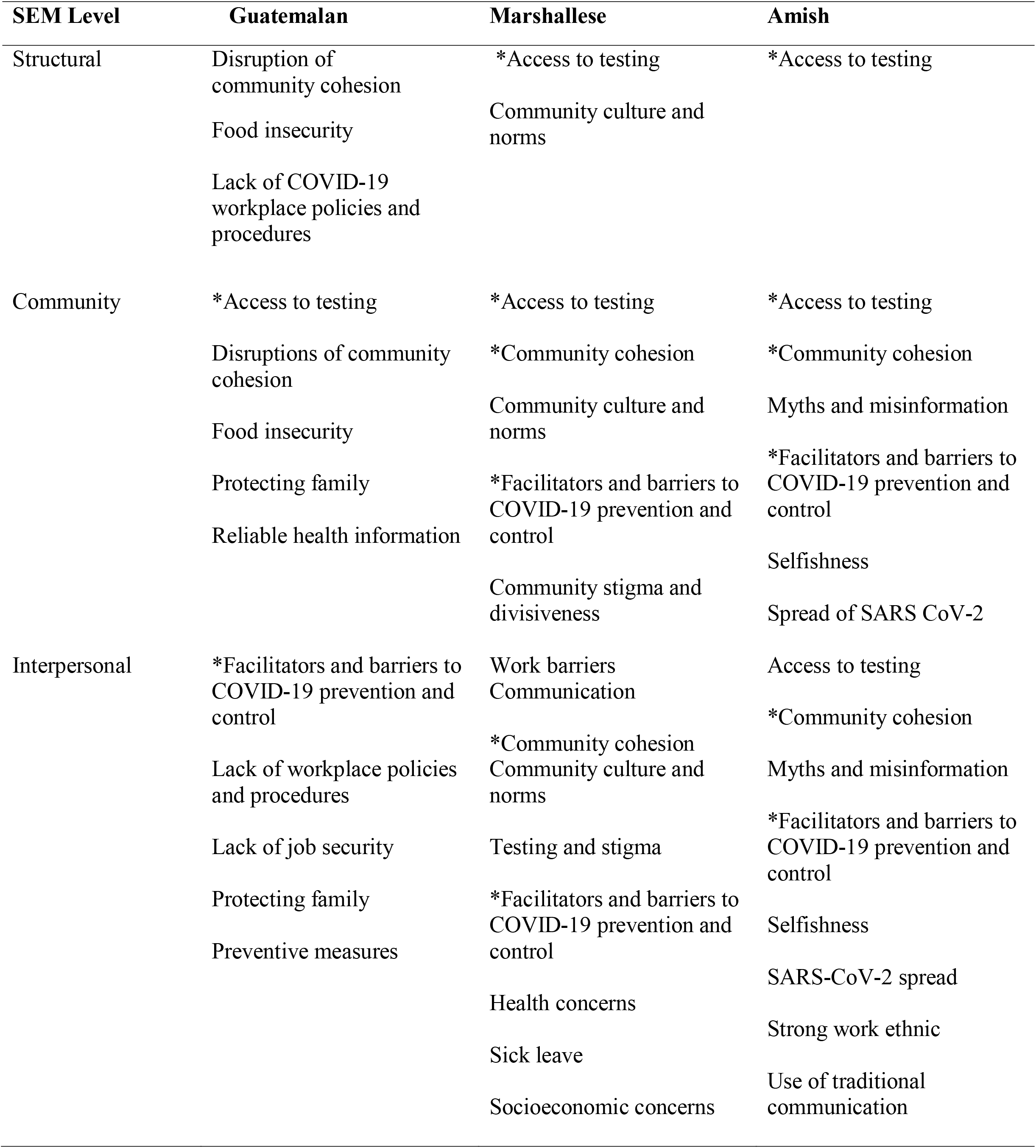
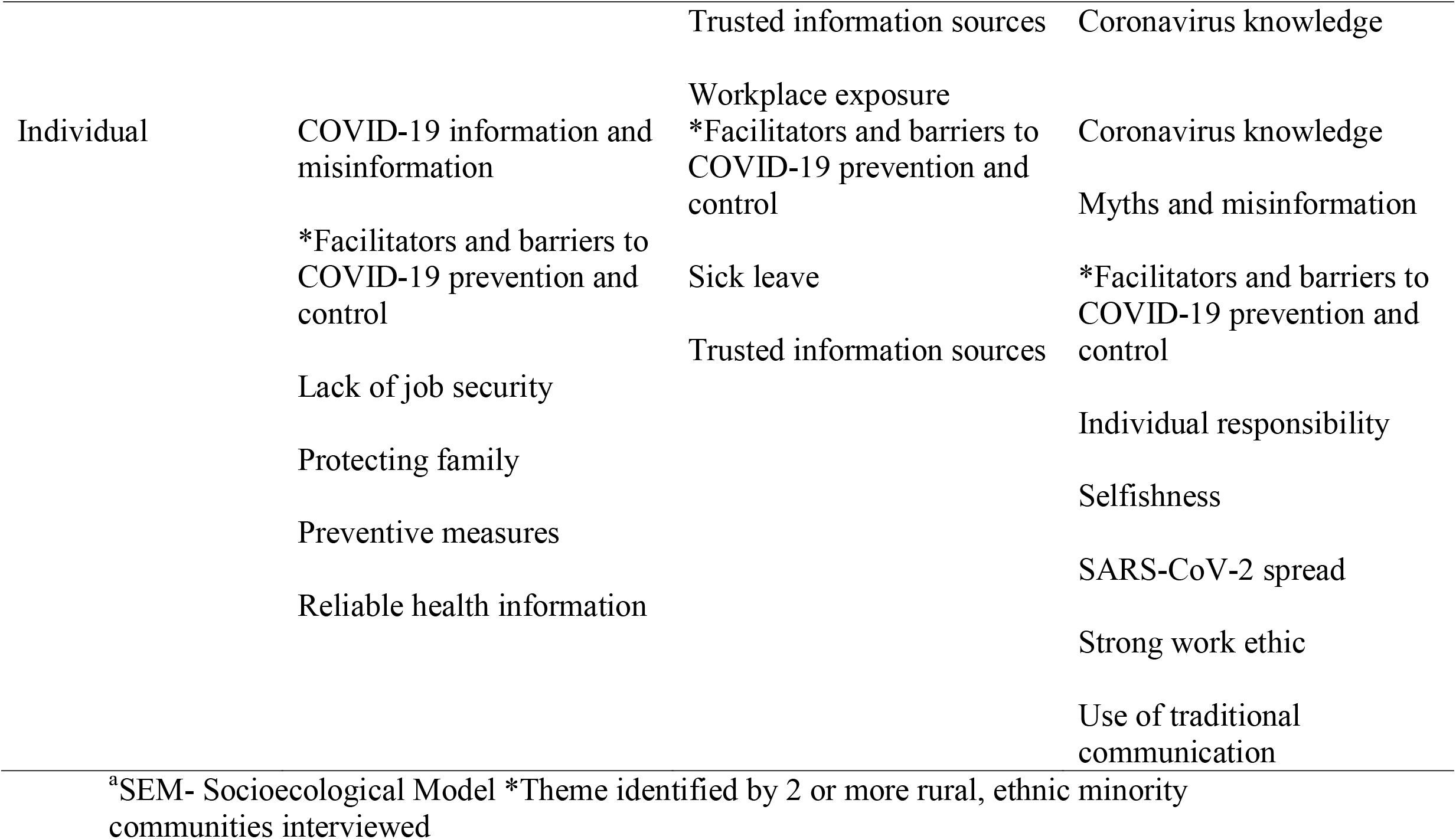
COVID-19 Themes Categorized by SEM^a^ Levels and Ethnic Minority Group

### Themes: Guatemalan community members and leaders

The study team in Ohio collaborated with a non-profit community organization to gain access to Guatemalan community leaders and members. Participants included meat processing plant, farm, and other food industry employees (n =5), faith-based community leaders (n=2), legal assistants for a community-based organization (n=2), a construction worker (n=1), a store cashier/community-based immigration advocate (n=1), and a self-employed community member (n=1). Investigators identified communication barriers, facilitators and barriers to COVID-19 prevention and control, food insecurity, lack of workplace safety policies, and COVID-19 knowledge and misinformation as overarching themes. One respondent shared that communication barriers contributed to lack of knowledge and risk of increased person-to-person transmission, as *“many of the families don’t speak the language and many places don’t have bilingual signs…lack of clear communication is [a] reason why it is difficult*.*”*

Another community member added that *“some people don’t believe, and some don’t care…some people don’t understand English, and some don’t understand Spanish. They don’t believe they are at risk to get the virus*.*”* COVID-19 knowledge varied among respondents. A community member shared that *“…you can get it from someone who is infected. Being around a person who has it and if you visit that person maybe you can get it*.*”* Lack of COVID-19 information, knowledge, and perception of risk were cited as barriers to infection prevention and control within Guatemalan communities and were grouped under the COVID-19 knowledge and misinformation theme. A community member shared that *“…they don’t care, they don’t want to follow orders even if [it is] in their best interest. Until they know somebody close gets it…they act when it’s too late*.*”* Another interviewee noted that *“perhaps the necessary measures have not been taken. Perhaps we do not have as much knowledge about the disease*.*”*

When asked specifically about facilitators for and barriers to COVID-19 prevention and control, including the ability to practice social distancing, an interviewee who had COVID-19 several weeks before the interview shared that *“It’s hard to pretty much be away from my kids. They are attached to me. I couldn’t hug them for 14 days…was hard*.*”* Additionally, an interviewee expressed frustration with community members who did not adhere to recommended preventive measures such as mask wearing and social distancing when she stressed that “*in the stores or in the streets, you try to follow social distancing suggestions…it makes it hard when not everybody believes it*.*”*

Some community members reported that multiple families resided in one home *“to share the rent, to share the cost. If one person is infected, [there is] no place to isolate*.*”* For some community members, sharing a home with multiple families was a barrier to quarantining and isolating successfully to reduce the spread of COVID-19, but was a necessity due to economic constraints. Several community members experienced food insecurity during the pandemic, as the *“local food pantry shut down. That was a big hit. With them shutting down and people not working, people were struggling a lot*.*”*

One respondent described that *“…when I was sick, my family were afraid to go out and we have kids. They had milk, but we didn’t have enough food*.*”* Several respondents mentioned lack of workplace safety policies as a barrier to preventing SARS-CoV-2 transmission due to *“lack of information, lack of care from employers to provide a safe environment and safe workplace for employees*.*”* A community member who recovered from COVID-19 added that *“I stayed home for 3 weeks [and] they didn’t send me a check. They don’t care if you are sick, they start calling [you] to go back to work or you could lose your job*.*”*

### Themes: Marshallese community members and leaders

The county health department in Ohio, meat and poultry processing and packing plants, and community leaders worked closely with investigators to arrange interviews with Marshallese community members. The study team interviewed local health department staff (n=3), meat processing plant managers (n=2), and Marshallese community members (n=2). Prominent themes included facilitators and barriers to COVID-19 prevention and control, stigma, and community culture and norms. When asked about facilitators for and barriers to COVID-19 prevention and control, a county health department staff member highlighted the reality of crowded living conditions for some families by noting that “*apartments are supposed to be for 6 people, really you find that they are living with 10 people. It’s hard for public health*.” Stigma was also mentioned as an ongoing fear among community members in that *“there’s some prejudice against the Marshallese…they are linked to the outbreak*.*”*

Additionally, one interviewee shared that there’s a *“stigma that goes with the Islander population…stigma with landlords…if more people are living together [than] should be or allowed…there’s a lot of stigma around it*.*”* Several participants identified community barriers to routine use of face masks and adoption of social distancing as a regular practice among community members, as *“some people look at it as infringement on their freedom and people don’t believe it [COVID-19] is here and they are tired of following the rules*.*”* Investigators learned that some Marshallese community members experienced stigma inside and outside of their community because of getting tested for COVID-19, as one respondent stressed that *“my concerns are about blame…*.*if you go get a test, then there is stigma”* and *“they [Marshallese] are afraid to take a test because they are labeled*.*”*

Similar to previous infectious disease outbreak investigations, we observed that socializing, getting together frequently, and being community-focused are normal practices ingrained in Marshallese culture that create and reinforce strong, close-knit communities (19-20). A community member shared that *“it’s natural to gather together…in our culture, [it’s] normal”* and *“I told everyone at my church not to gather and stay at home*.*”* Socioeconomic concerns were also mentioned as a barrier to social distancing and staying home when sick, as *“company ABC will send you home for 14 days and they don’t pay you*.*”*

### Themes: Amish community members and leaders

We interviewed Amish community members and leaders from two rural Ohio counties. Participants consisted of local business owners (n=2), faith-based and community leaders (n=3), a company manager (n=1), self-employed community members (n=2), a retired community member (n=1), and homemakers (n=2). Major themes that emerged from interviews included COVID-19 knowledge, misinformation and myths, community cohesion, and facilitators for and barriers to COVID-19 prevention and control. Knowledge varied among community members, but most respondents possessed some understanding about how SARS-CoV-2 was transmitted and how to prevent passing the virus on to others. A respondent mentioned that *“I think it spreads through physical contact. Handshaking, getting close. Interacting with people who have symptoms or are at risk for virus. Again, it is, I think the social gathering, [social] functions*.*”*

Moreover, a participant shared that SARS-CoV-2 could be transmitted through *“coughing, sneezing, through droplets, [and] touching [your] face…mask is more to protect you from touching your face*.*”* Another community member who tested negative for COVID-19 added that *“I find it as a “hidden monster”-very difficult to identify. I know a few of the symptoms*…*it lasts a couple of days*.” Additional interview participants described methods to prevent person-to-person transmission by *“…handwashing, social distancing, and staying at home,”* and having *“no physical contact with people outside the home…definitely stay home if you’re not feeling well*.*”* Members of Amish communities shared that it was challenging to use face masks regularly and practice social distancing.

Referring to social distancing, one community member said that “*we don’t practice it. I guess we don’t find it necessary*.” When asked about mask use to prevent COVID-19, an interviewee stated that “*what makes it hard for me is that mask use makes [my] glasses fog up and mask falls down; doesn’t sit right on my face*.” We also learned from community members that misinformation and myths circulated throughout the community such as *“far-fetched rumors about the virus…[like] some think there is a chip in vaccination that tracks people if you get vaccine*.*”* Another community member noted that there are *“a lot of rumors about the virus…[The] bible tells us about the “Mark of the Beast*.*”*

There seemed to be a reliance on natural remedies in the community, as an interviewee shared that *“we aim to drink a lot of water, take vitamin C, and immune boosters”* to stay healthy and protected from the virus. A few participants also spoke about being at low risk for COVID-19 and mentioned *that “I don’t think [the] risk is high to get it”* and that *“It is in the big towns, not among the [Amish]*.*”* Community cohesion is a significant part of Amish culture, which is reinforced and facilitated via regular social connection (21). Amish communities use social occasions (e.g., weddings, baptisms, funerals) and spiritual activities (church attendance) to connect with friends and family members.

A participant who recuperated from COVID-19 felt supported by his community and shared that the *“Bishop checked on me every day to see how I was doing. I had a big circle of friends that called us every day*.*”* Another community member emphasized the importance of family, as he shared that a “*sense of family and connection with family has more healing power. We understate [the] value to be able to connect with family*.*”* A community leader added that *“fellowship is as important to us as worship”* while another participant said that *“to meet each other with a handshake in the morning…[it’s] culturally ingrained in our way of having services*.*”*

As we interviewed members of the Amish community, we learned that several leaders were open to collaborating with the local health department and CDC to disseminate critical health education messages using a variety of local communication mediums (e.g., Amish newspaper, radio, magazine, health information briefs) to quell SARS-CoV-2 transmission and reinforce simple preventive measures.

## Discussion

Overall, we found evidence of health inequities among the ethnic minority groups we interviewed in rural Ohio. Health seeking behavior was influenced by health care access, potential for experiencing stigma from the community, and fear of police involvement. Meeting basic family needs such as paying rent, buying supplies, and acquiring groceries was challenging for some of the Guatemalan and Marshallese community members we interviewed. Participants from all three communities mentioned the sharing of myths and misinformation about COVID-19 within the community, which underscores the importance of disseminating simple, timely, and culturally and linguistically appropriate health information to all communities through trusted leaders.

Additionally, in some communities, we learned from community members and leaders that COVID-19 testing events would be most successful and have the greatest reach if organized and implemented through community-based organizations in collaboration with local health departments. Results from qualitative interviews showed significant gaps in COVID-19 knowledge and awareness, challenges with workplace policies and procedures addressing worker safety and compensation, reliable health information, health care access, and challenges with community cohesion when implementing COVID-19 mitigation strategies. Marshallese and Guatemalan respondents reported fear, stigma, and economic hardship as barriers to COVID-19 prevention and control. Food insecurity was identified as a barrier during the COVID-19 pandemic among participants from Guatemala residing in rural Ohio.

Knowledge about COVID-19 transmission and prevention varied among participants from the Amish communities. All participants identified the use of masks and social distancing as the most difficult preventive measures to adopt due to lack of comfort, practicality, cultural acceptability, and convenience. There is a higher risk of COVID-19 in congregate settings, including among closely-knit communities due to cultural practices, lack of COVID-19 knowledge, certain community and personal beliefs, and economic hardship (12, 22). As a result of our interviews with community members representing three ethnic groups, we found that disease outbreaks occurred due to living conditions, cultural norms and practices, and misperceptions about coronavirus and transmission risk. Strategies to reduce SARS-CoV-2 transmission include routine collaboration among health departments, community-based organizations, and community leaders, which promotes public health knowledge, situation awareness, and resource sharing. This also strengthens community partnerships, interest, and trust.

This type of collaboration is imperative during planning and implementation of COVID-19 testing events, community initiatives to reduce food insecurity, and COVID-19 health education efforts. This ongoing relationship builds a robust foundation for an integrated, informed, and collective response to public health threats like COVID-19 and other infectious disease outbreaks. Consistency that is built over years with active participation and buy-in from community members creates a unified approach to infectious disease prevention and response that is feasible and sustainable. Respondents consistently indicated that tailored health education messages that included appropriate and culturally acceptable language would be better received by their community members.

Communication and COVID-19 knowledge and awareness (e.g., SARS-CoV-2 transmission, COVID-19 prevention measures, COVID-19 symptoms, knowledge about COVID-19 disease as a pandemic, SARS-CoV-2 testing) barriers existed in all three groups we interviewed. Timely, clear, and consistent public health messaging about prevention and control activities, tailored for the intended audience, could help address these barriers. For example, use of the terms physical distancing instead of social distancing could be more acceptable in these communities due to the importance of family and community cohesion.

For close-knit communities, where family ties are very strong, COVID-19 prevention messages may focus on family importance and keeping family members healthy, especially for those who go to work as well as messaging around COVID-19’s potential impact on mental and social health within families. Messages should be shared using multiple outlets and trusted community leaders. Consistent and ongoing community sensitization on major communicable disease risks, including prevention messages and active community engagement (e.g., through community events, print and social media, workplace and school-based education, training, and community-driven learning initiatives) could facilitate community acceptance and adoption of public health recommendations.

There were several strengths to our study approach. First, we focused on the perspectives of both community members and leaders to understand how COVID-19 impacted these communities at the individual, interpersonal, community, and structural levels. This multi-faceted approach based on the SEM allowed us to understand facilitators for and barriers to COVID-19 prevention and control, community norms, and community needs among three ethnic minority communities residing in rural areas of Ohio. Second, we partnered with state and county health departments as well as community-based organizations to gain access to the communities, which helped us complete the interviews in a short two-week timeframe.

## Limitations

Our study findings are subject to some limitations. First, our focus was on three ethnic minority communities residing in rural Ohio, and we were only able to interview a few Marshallese community members and leaders. Therefore, our study findings may not be generalizable to all members of these groups and are likely not generalizable to the United States. Additionally, some of the interviews were conducted by phone due to the short data collection timeframe and for convenience, participant privacy, and safety reasons. Lastly, interviews with some Guatemalan community leaders and members relied on the skills of the interpreter or translator. Resulting interview notes may not have fully captured the original intent of interviewee responses.

### Lessons Learned

Collaboration with community-based organizations (CBOs) was instrumental in gaining entry and access to the communities due to the CBO credibility and trust within the community. Routine health department engagement with, and feedback from, CBOs can fuel community buy-in, which can lead to meaningful and lasting partnerships to prevent disease and promote health equity. Interest, effort, and time invested in Amish communities to establish and maintain ongoing collaborations may promote health and prevent disease. Learning from previous culturally appropriate infectious disease response activities (e.g., measles vaccination in response to community outbreaks) may provide insight for tailoring COVID-19 health education, testing, and outreach to Amish populations (23).

Understanding population-specific barriers and providing necessary resources, healthcare services, education, and social support tailored to each population could help reduce further SARS-CoV-2 transmission within unique communities. This will be especially important, for example, for the reach and success of COVID-19 immunization campaigns during vaccine rollout. To help support sustainability and ensure a lasting impact, piloting interventions to assess their acceptability and uptake by community members is a critical step before implementation occurs at individual, family, community, organizational, and structural levels.

## Data Availability

All data produced in the present work are contained in the manuscript.

## Acknowledgments

The authors wish to thank the community-based organizations, county and local health departments, and rural community members in Ohio who graciously shared their time, perspectives, and experiences with us.

## Footnotes

i See e.g., 45 C.F.R. part 46.102(l)(2); 21 C.F.R. part 56; 42 U.S.C. §241(d); 5 U.S.C. §552a; 44 U.S.C. §3501 et seq.

## References

1. Bialek, S, Boundy, E, Bowen, V, et al. (2020) CDC COVID-19 Response Team. Severe outcomes among patients with coronavirus disease 2019 (COVID-19)—United States, February 12–March 16, 2020. MMWR Morb Mortal Wkly Rep, 69:343–346. https://doi.org/10.15585/mmwr.mm6912e.

2. Centers for Disease Control and Prevention. (2020, August 24) Coronavirus Disease 2019. If you are immunocompromised, protect yourself from COVID-19. https://www.cdc.gov/coronavirus/2019-ncov/need-extra-precautions/immunocompromised.html.

3. Moore, J, Ricaldi, J, Rose, C, et al. (2020) Disparities in incidence of COVID-19 among underrepresented racial/ethnic Groups in counties identified as hotspots during June 5– 18, 2020 — 22 States, February–June 2020. MMWR Morb Mortal Wkly Rep, 69: 1122– 1126. doi: http://dx.doi.org/10.15585/mmwr.mm6933e1

4. Self, W, Tenforde, M, Stubblefield, W, et al. (2020) Seroprevalence of SARS-CoV-2 among frontline health care personnel in a multistate hospital network — 13 academic medical centers, April–June 2020. MMWR Morb Mortal Wkly Rep, 69:1–7. doi: http://dx.doi.org/10.15585/mmwr.mm6935e2

5. Centers for Disease Control and Prevention. (2021, March 4) Coronavirus disease 2019. Cases in the U.S. 2020. https://www.cdc.gov/coronavirus/2019-ncov/cases-updates/cases-in-us.html.

6. Ohio Department of Health. (2021, March 4) Coronavirus disease 2019. https://coronavirus.ohio.gov/wps/portal/gov/covid-19/home

7. Cafer, A, Rosenthal, M. (2020, July 20) COVID-19 in the rural south: A perfect storm of disease, health access, and co-morbidity. APCRL Policy Briefs. 2. https://egrove.olemiss.edu/apcrl_policybriefs/2.

8. Paul, R, Arif, A, Adeyemi, O, Ghosh, S, Han, D. (2020) Progression of COVID□19 from urban to rural areas in the United States: A spatiotemporal analysis of prevalence rates. J Rural Health, 00:1–11. doi: 10.1111/jrh.12486

9. Sood, L, Sood, V. (2020) Being African American and rural: A double jeopardy from COVID□19. J Rural Health, 0:1–5. doi:10.1111/jrh.12459.10

10. Stuesse, A, Dollar, N. (2020, September 24) Economic Policy Institute. Working economics blog. Who are America’s meat and poultry workers? https://www.epi.org/blog/meat-and-poultry-worker-demographics/

11. Dyal, J, Grant, M, Broadwater, K Bjork, et al. (2020) COVID-19 among workers in meat and poultry processing facilities-19 states. MMWR Morb Mortal Wkly Rep, 69(18):557–561.

12. Lessler J, Azman AS, McKay HS, Moore SM. (2017) What is a Hotspot Anyway? Am J Trop Med Hyg, 96(6):1270–1273. doi:10.4269/ajtmh.16-0427

13. McLeroy, K R, Bibeau, D, Steckler, A Glanz, (1988) K. An ecological perspective on health promotion programs. Health Educ Q, 15(4):351–377. https://doi.org/10.1177/109019818801500401

14. Frieden, T. (2010) A framework for public health action: the health impact pyramid. Am J Public Health, 100(4):590–595. doi:10.2105/AJPH.2009.185652

15. Dowling, M. (2007) From Husserl to van Manen. A review of different phenomenological approaches. Int J Nurs Stud, 44(1):131-142. 10.1016/j.nurstu.2005.11.026

16. Husserl, E. (2012) Ideas: General Introduction to Pure Phenomenology. Routledge.

17. Moutsakas, C. (1994) Phenomenological Research Methods (2nd Ed.). Sage. Retrieved October 19, 2020 from https://dx.doi.org/10.4135/9781412995658

18. Mason, J. (2002) Qualitative Researching. Sage.

19. Fields. V, Safi, H, Waters, C., et al. (2019) Mumps in a highly vaccinated Marshallese community in Arkansas, USA: an outbreak report. Lancet Infect Dis, 19(2):185–192. doi:10.1016/S1473-3099(18)30607-8.

20. Marx, G, Burakoff, A, Barnes, M, et al. (2018) Mumps outbreak in a Marshallese community-Denver metropolitan area, Colorado, 2016-2017. MMWR Morb Mortal Wkly Rep, 67(41):1143–46. http://dx.doi.org/10.15585/mmwr.mm6741a2

21. Hostetler, J. (1993) Amish Society (4^th^ Ed.). JHU Press.

22. James, A, Eagle Phillips, C., et al. (2020) High COVID-19 attack rate among attendees at events at a church-Arkansas, March 2020. MMWR Morb Mortal Wkly Rep, 69(20):632–635.

23. Gastañaduy, P, Budd, J, Fisher, N., et al. (2016) A measles outbreak in an underimmunized Amish community in Ohio. N Engl J Med, 375:1343–1354. doi: 10.1056/NEJMoa1602295

